# Oral health literacy, oral health behavior, and oral health status among dental patients and associated factors

**DOI:** 10.1101/2022.09.09.22279289

**Authors:** Ran An, Qianqian Li, Yuan Luo, Wenfeng Chen, Muhammad Sohaib, Meizi Liu, Zitong Wu

**Affiliations:** Xiangya School of Nursing, Central South University, Changsha, Hunan Province, China; The first Mobile Corps Hospital of the Chinese People’s Armed Police Force, Baoding City, Hebei Province, China; Teaching and Research Section of Clinical Nursing, Xiangya Hospital Central South University, Changsha, Hunan Province, China; Xiangya School of Nursing, Central South University; Children hospital and Institute of child health, Multan, Pakistan; Xiangya School of Nursing, Central South University, No.87 Xiangya Road, Kaifu District, Changsha, Hunan Province, China

**Keywords:** Oral health literacy, Oral health behavior, Oral health status, dental patients, Cross-sectional study

## Abstract

**Background:** Oral health plays an important role in overall health. Little is known about oral health literacy, oral health behavior, and oral health status in dental patients.

**Purpose:** This study aimed to assess oral health literacy (OHL), oral health behavior, oral health status, and associated factors in dental patients.

**Methods:** This cross-sectional study was conducted between June,13, 2022 and July, 26, 2022 in a tertiary general hospital, in Hebei, China. A total of 184 patients participated in the study. OHL was assessed by the Chinese version of the Health Literacy Dental Scale (HeLD-14). Trained interviewers performed face-to-face interviews for oral health-related behavior information. Data were analyzed using SPSS version 24. Mann–Whitney U-tests, chi-square tests, and binary logistic regression analysis were performed.

**Results:** Participants were 184 dental patients which consisted of 78.3% men and a mean age of 28.36 ± 10.72 years old. The mean oral health literacy score was 43.07±9.920 (out of 56). There were significant associations between inadequate OHL and economic burden (odds ratio [OR]=2.636, P = 0.003), mouthwash (OR=2.433, P = 0.006), gum bleeding (OR=3.798, P = 0.030), and dental visiting (OR=1.885, P = 0.049).

**Conclusion:** The oral health literacy of dental patients is at a medium level overall. Patients with inadequate OHL had a higher frequency of bleeding gums and less frequent dental visiting. Health care providers should consider improving oral health literacy among dental patients.

## Background

Globally, about 3.5 billion people worldwide suffer from oral diseases[1], oral diseases lead to serious health and economic burdens, causing pain, and sepsis, greatly reducing the quality of life [1]. The World Health Organization advocates the promotion of oral health care and public health practices and approved a resolution on oral health in 2021 to encourage the proactive promotion of one’s health [2, 3]. China has also introduced guidelines and policies such as the “Healthy China Action (2019-2030)” and the “Healthy Mouth Action Program (2019-2025)” to strengthen oral health management and oral education[4].

At present, studies on oral health research are mainly focused on teenagers, the elderly, pregnant women, and adults [5-7], while the literature on oral health and oral health behaviors of dental patients is sparse, who have a high prevalence of oral diseases, deserve more attention. According to the Fourth National Oral Health Survey (2015-2016) in mainland China[8], a large proportion (52.8%∼69.3%) of Chinese adults have periodontal disease [8]. In contrast to the high prevalence, less than 20% of Chinese adults were knowledgeable about periodontal disease. Moreover, little is known about the problems with their oral health behaviors and how they are affected by oral health problems and the associated factors.

In recent years, oral health literacy (OHL) has emerged as a possible underlying mechanism that explains oral health status in many groups [16-19], oral health literacy is the capacity to understand oral health information and use it to inform beneficial oral health prevention and treatment decisions [20]. OHL is regarded as being essential to the promotion of oral health and the avoidance of oral diseases [21]. Besides, OHL is regarded to largely reflect underlying social and personal values related to dental care, and oral health outcomes[9, 10]. A study in Iran showed that 62.5% of adult patients had an adequate oral health literacy level[9]. However, a survey of older adults in Thailand showed the majority of participants (85%) had inadequate functional health literacy,[11]. Previous studies show that people with limited OHL levels had poor periodontal health[12]. Oral health literacy has also been confirmed to be strongly associated with oral health behaviors[13] and oral health-related quality of life[14].

To the best of our knowledge, no prior research was conducted in China to evaluate oral health literacy in dental patients. The purpose of this study is to evaluate oral health literacy, oral health behaviors, and oral health status in dental patients and the associated factors, which are important for improving oral health status and quality of life.

## Methods

### Study design

A descriptive cross-sectional study design was conducted from June, 13, 2022 to July, 26, 2022 with 184 patients aged from 18 to 77 years old that attended the Department of Stomatology of the First Mobile General Hospital of Armed Police, Hebei, China.

### Inclusion and exclusion criteria

1. Inclusion criteria: aged ⩾ 18 years, who attended at the dental clinic; clear consciousness, able to make independent judgments and complete the questionnaire; informed consent and voluntary participation in the study
2. Exclusion criteria: mental disorders or other serious illnesses, unable to cooperate with the completion of the questionnaire.

### Sampling

Since there is no survey on oral health literacy for outpatients in the dental clinic, we refer to the survey on oral health literacy of adults[6], whose incidence of low level of oral health literacy is about 12.4%, n=u_2_ɑp(1-p)/δ^2^, so this study takes p=0.124 as the basis for estimating the sample size, n = the desired sample size when the population is greater than 10,000,δ is the allowable error, δ=0.05 is set in this study, take α= 0.05, u= the standard normal deviate, usually set at 1.96 which corresponds to 95% confidence level, the final calculation of the sample content required for this study n≈ 162, the effective response rate is calculated according to 90%, then the sample size required for this study is 180.

### Data collection

Data were collected by 2 trained nurses who had prior experience with national survey data collection. The nurses met with potential participants to explain the aim of the study and the process of data collection. Those willing to participate were asked to complete an informed consent form to the researcher. Participants were asked to complete the questionnaire on their own without interruption, which consisted of a socio-demographics questionnaire and an oral health literacy scale.

### Socio-demographics questionnaire

A self-administered structured questionnaire was adapted after reviewing the relevant literature[15-17]. It had been validated and pretested on 30 health professionals working at the hospital for its consistency. The questionnaire has three main parts: (1) The first part contains items on sociodemographic information of participants, such as age, gender, place of residence, marital status, whether living alone, education level, smoking history, drinking history, sleep status and monthly household incomes; (2) The second part is related to participants’ information on clinical characteristics, such as the reason for consultation and disease type; (3) The third part contains 7 items to evaluate oral health behaviors and oral health status, including daily brushing frequency, daily rinsing, whether mouthwash daily, flossing, whether there was gum bleeding when brushing, whether they had dental visiting in the past year, learning bout oral health knowledge, and self-assessment of oral health status.

### Oral Health Literacy

The simplified version of the Health Literacy Dental Scale -14(HeLD-14) was developed by Jones[18]based on the full version of the Health Literacy Dental Scale (HeLD-29). Chinese scholar Yan[19] adapted and tested the Chinese version of the scale, which is a valid and reliable questionnaire for this purpose, before the study started, we obtained the authorization of the authors. The Chinese version of the scale’s Cronbach’s alpha value was 0.89, and the test-retest reliability was 0.786, indicating an adequate level of inter-item reliability. The scale has 14 questions divided equally into seven dimensions. Each item was scored using a 5-point Likert scale ranging from 1 (without any difficulty) to 5 (unable to do). The analysis first converted the 5 points to 0, the 4 points to 1, the 3 points to 2, the 2 points to 3, the 1 point to 4, then the total score was calculated, with higher scores indicating higher oral health literacy.

### Statistical analysis

The data were analyzed using SPSS statistical software version 24.0 (SPSS, Central south university, China). Descriptive statistics were used to describe the demographic characteristics and general information among patients. Associations of the OHL with general demographic information, clinical characteristics information, and oral health behaviors were analyzed by Chi-square tests. Based on the median split, HeLD-14 scores were dichotomized into two groups (⩾ 44 vs.<44), and a binary logistic regression model was used for the analysis. A multivariate logistic regression model was developed to test associations between OHL and other potential associated factors. Differences were considered statistically significant at *P*<0.05.

### Ethical approval and informed consent

The study was performed by the Declaration of Helsinki. All respondents gave informed consent prior to conducting the interview. The study protocol was approved by the College of Nursing, Central South University, Nursing and Behavioral Medicine Research Ethics Review Committee (E202297).

## Results

In total, 200 questionnaires were distributed and 188 copies were collected. After checking and excluding 4 cases of invalid questionnaires, 184 valid questionnaires were recovered, with a valid recovery rate of 92%. Among the 184 individuals included in the analysis, the mean age was 28.36 ± 10.72 (SD), and most of the patients were male (78.3%), had at least senior secondary education (92.4%), unmarried (61.9%), and not currently smoking (60.3%). The reasons for dental visiting were mainly swollen or bleeding gums (50.5%), about 34.8% of them were diagnosed with periodontitis. About 45.7% of the sample attended dental appointments in the last year, and more than half of them (69.6%) self-assessed oral health status as fair.

The patients’ OHL score levels ranged from 4 to 56 (43.07±9.920), and the overall scores were normally distributed. A total of 46.7% of the study participants had adequate OHL with a median HeLD score of 44, we then used this number (44) to categorize the study subjects into inadequate OHL and adequate OHL groups. Participants who had HeLD scores ⩽ 44 inadequate OHL, and those with HeLD scores > 44 were categorized as adequate OHL.

Table 1 shows the common characteristics associated with oral health literacy. Significant differences in OHL scores were noted between different BMI classifications, sleep status, economic burden, whether they had seen a dentist in the last year, whether to rinse their mouth, dental Scaling, and whether their gums bled while brushing.

**Table 1.** Comparison of Oral health literacy scores among patients with different demographic characteristics (N = No. of participants, % = percent)

| Variables | Total<br>(%) | Inadequate<br>OHL | High OHL | $\chi^2$<br>value | *P*<br>value |
| --- | --- | --- | --- | --- | --- |
| **Gender** |  |  |  | 0.705 | 0.401 |
| Male | 144(78.3) | 72(75.8) | 72(80.9) |  |  |
| Female | 40(21.7) | 23(24.2) | 17(19.1) |  |  |
| **Ethnicity** |  |  |  | 0.008 | 0.930 |
| Han Chinese | 177(96.2) | 92(96.8) | 85(95.5) |  |  |
| Ethnic Minority | 7(3.8) | 3(3.2) | 4(4.5) |  |  |
| **Age group(years)** |  |  |  | 6.672 | 0.083 |
| 18~25 | 104(56.5) | 56(58.9) | 48(53.9) |  |  |
| 26~35 | 54(29.3) | 23(24.2) | 31(34.8) |  |  |
| 36~55 | 18(9.8) | 9(9.5) | 9(10.1) |  |  |
| ≥56 | 8(4.3) |  |  |  |  |
| **BMI classification** |  |  |  | 8.359 | **0.015** |
| Underweight | 6(3.3) | 6(6.3) | 0(0.0) |  |  |
| Normal weight | 123(66.8) | 60(63.2) | 63(70.8) |  |  |
| Overweight | 55(29.9) | 29(30.5) | 26(29.2) |  |  |
| **Education level** |  |  |  | 3.766 | 0.439 |
| Primary | 7(3.8) | 5(5.3) | 2(2.2) |  |  |
| Junior Secondary | 7(3.8) | 5(5.3) | 2(2.2) |  |  |
| Senior Secondary | 41(22.3) | 23(24.2) | 18(20.2) |  |  |
| College | 69(37.5) | 31(32.6) | 38(42.7) |  |  |
| Bachelor's and above | 60(32.6) | 31(32.6) | 29(32.6) |  |  |
| **Smoking** |  |  |  | 0.156 | 0.693 |
| Non-smoker | 111(60.3) | 56(58.9) | 55(61.8) |  |  |
| Smoker | 73(39.7) | 39(41.1) | 34(38.2) |  |  |
| **Alcohol consumption** |  |  |  | 0.704 | 0.402 |
| No | 129(70.1) | 64(67.4) | 65(73.0) |  |  |
| Yes | 55(29.9) | 31(32.6) | 24(27.0) |  |  |
| **Work status** |  |  |  | 4.533 | 0.209 |
| Full-time | 75(40.8) | 42(44.2) | 33(37.1) |  |  |
| Unemployed | 16(8.7) | 11(11.6) | 5(5.6) |  |  |
| Retired | 5(2.7) | 3(3.2) | 2(2.2) |  |  |
| Other | 88(47.8) | 39(41.1) | 49(55.1) |  |  |
| **Marital Status** |  |  |  | 1.992 | 0.158 |
| Married | 71(38.6) | 32(33.7) | 39(43.8) |  |  |
| Unmarried | 112(61.9) | 63(66.3) | 50(56.2) |  |  |
| **Living alone** |  |  |  | 0.705 | 0.401 |
| Yes | 144(78.3) | 72(75.8) | 72(80.9) |  |  |
| No | 40(21.7) | 23(24.2) | 17(19.1) |  |  |
| **Sleeping status** |  |  |  | 6.904 | **0.032** |
| Good | 115(62.5) | 53(55.8) | 62(69.7) |  |  |
| Fair | 61(33.2) | 35(36.8) | 26(29.2) |  |  |
| Poor | 8(4.3) | 7(7.4) | 1(1.1) |  |  |
| **Place of residence** |  |  |  | 0.870 | 0.351 |
| Urban | 122(66.3) | 60(63.2) | 62(69.7) |  |  |
| Rural | 62(33.7) | 35(36.8) | 27(30.3) |  |  |
| **Monthly household income (RMB)** |  |  |  | 4.302 | 0.231 |
| ≤2000 | 19(10.3) | 12(12.6) | 7(7.9) |  |  |
| 2000~5000 | 36(19.6) | 22(23.2) | 14(15.7) |  |  |
| 5000~10000 | 94(51.1) | 47(49.7) | 47(52.8) |  |  |
| ≥10,000 | 35(19.0) | 14(14.7) | 21(23.6) |  |  |
| **Economic burden** |  |  |  | 11.826 | **0.008** |
| None | 71(38.6) | 26(27.4) | 45(50.6) |  |  |
| Light | 30(16.3) | 17(17.9) | 13(14.6) |  |  |
| Fair | 69(37.5) | 45(47.4) | 24(27.0) |  |  |
| Heavy | 14(7.6) | 7(7.4) | 7(7.4) |  |  |
| **Reason for dental visiting** |  |  |  | 1.830 | 0.401 |
| Swollen or bleeding gums | 93(50.5) | 44(46.3) | 49(55.1) |  |  |
| Other discomforts | 58(31.5) | 34(35.8) | 24(27.0) |  |  |
| Check-up or consultation | 33(17.9) | 17(17.9) | 16(18.0) |  |  |
| **Disease Type** |  |  |  | 4.749 | 0.461 |
| Caries | 32(17.4) | 18(18.9) | 14(15.7) |  |  |
| Periodontitis | 64(34.8) | 28(29.5) | 36(40.4) |  |  |
| Oral mucosal disease | 4(2.2) | 3(3.2) | 1(1.1) |  |  |
| Missing teeth | 19(10.3) | 12(12.6) | 7(7.9) |  |  |
| Fractured tooth | 4(2.2) | 3(3.2) | 1(1.1) |  |  |
| Other | 61(33.2) | 31(32.6) | 30(33.7) |  |  |
| **Daily brushing frequency(times)** |  |  |  | 0.516 | 0.773 |
| 1 | 28(15.2) | 16(16.8) | 12(13.5) |  |  |
| 2 | 143(77.7) | 73(76.8) | 70(78.7) |  |  |
| ≥3 | 13(7.1) | 6(6.3) | 7(7.9) |  |  |
| **Mouth Rinse daily** |  |  |  | 4.790 | 0.029 |
| No | 86(46.7) | 49(55.1) | 37(38.9) |  |  |
| Yes | 98(53.3) | 40(44.9) | 58(61.1) |  |  |
| **Dental cleaning** |  |  |  | 4.419 | **0.036** |
| No | 135(73.4) | 76(80.0) | 59(66.3) |  |  |
| Yes | 49(26.6) | 19(20.0) | 30(33.7) |  |  |
| **Gingival bleeding on brushing** |  |  |  | 7.302 | **0.007** |
| Never or sometimes | 163(88.6) | 82(95.3) | 81(82.7) |  |  |
| Often | 21(11.4) | 4(4.7) | 17(17.3) |  |  |
| **Visited the dentist in the past year** |  |  |  | 3.856 | 0.050 |
| Yes | 84(45.7) | 50(52.6) | 34(38.2) |  |  |
| No | 100(54.3) | 45(47.4) | 55(61.8) |  |  |
| **Self-rated oral health** |  |  |  | 3.446 | 0.178 |
| Poor | 28(15.2) | 16(16.8) | 12(13.5) |  |  |
| Fair | 128(69.6) | 69(72.6) | 59(66.3) |  |  |
| Good | 28(15.2) | 10(10.6) | 18(20.2) |  |  |

Table 2 presents the findings of the multivariable modelling by testing the independent variables associated with OHL. In the final model, the risk indicators included economic burden, mouthwash, gum bleeding, and dental visiting. Participants with economic burden tend to have inadequate OHL than their counterparts, and tend to be less likely to rinse their mouth daily or visit the dentist than people with OHL, and patients with inadequate OHL tend to have gum bleeding frequently.

**Table 2.** Logistic regression analysis of oral health literacy in dental patients

| Independent Variables | $\beta$ | SE | Wald<br>$\chi^2$ | P | OR | 95%CI |
| --- | --- | --- | --- | --- | --- | --- |
| Economic Burden | 0.969 | 0.331 | 8.577 | 0.003 | 2.636 | (1.378-5.044) |
| Mouthwash (1) | 0.889 | 0.326 | 7.445 | 0.006 | 2.433 | (1.285-4.608) |
| Gum bleeding | 1.335 | 0.616 | 4.700 | 0.030 | 3.798 | (1.137-12.694) |
| Dental visiting (1) | 0.634 | 0.322 | 3.879 | 0.049 | 1.885 | (1.003-3.543) |
| Constants | -1.175 | 0.339 | 12.036 | 0.001 | 0.309 |  |

## Discussion

In this study, we examined oral health literacy and its associations with socio-demographics, oral health behaviors, and self-assessed oral health status among dental patients in China. Our research shows that oral health literacy was significantly related to socio-demographics, oral health behaviors, and oral health status but not to self-assessed oral health.

The results of this study showed that the OHL score of 184 patients was (43.46 ± 9.239), which indicates that OHL was generally moderate in this sample. The score is in agreement with a study of older adults in Brazil[20] but is much lower than studies on adults in Brazil [14] and Malaysia [14] and Malaysia[21]. The reason may be the inequality in oral and dental health services in China, including social, economic, cultural, and environmental factors, insurance policies, and practices[22]. Moreover, there is a great imbalance in the distribution of dental practitioners among the provinces in China, neither the number of dental practitioners nor the number of dental visits is enough to meet the dental care demand in China as compared to developed countries[23]. There are also some personal factors, such as personal characteristics, health needs, and health behaviors[9, 11].

Among the reasons for consultation, more than half of the patients were due to gum swelling or bleeding (50.5%), other discomforts accounted for about one-third, and only 17.9% of the patients were due to physical examination or consultation, which is much lower than a study in the United States[21], which showed that more than half of the people (52.4%∼60.5%) took the initiative for physical examination or consultation, while only 6.3%∼7.6% of the people sought medical treatment for various discomforts, indicating that the awareness to seek oral health service utilization of patients in China is not good enough. The 4th National Oral Health Survey (2015-2016) in China indicate that about 20% of adults utilized oral health services in the past 12 months, but most of them (78.7%∼93.7%) visited a dentist for treatment, and most of them sought the service motivated by pain[24]. Unfortunately, a relatively low level of dental service utilization is typically associated with a greater economic burden[25]. Increasing awareness of the importance of preventative rather than symptomatic dental visits will help to improve oral health and optimize dental care support, in line with China’s longstanding national policy of a prevention-oriented model for the oral healthcare system and dental public health system[26].

Periodontitis (34.8%) and caries (17.4%) are common diseases among the patients who come to the clinic. Globally, dental caries and periodontal diseases (gingivitis and periodontitis) affect the majority of people, with treatment costs dominating the health care system[1]. The 4th National Oral Health Survey (2015-2016) in China shows that the percentage of periodontitis was about 52.8%∼64.6%[27]. Past studies linked periodontal disease with many other non-infectious systemic diseases[28], and a recent review suggests an association between periodontal disease and Alzheimer’s disease[29]. Periodontal disease is negatively correlated with oral health-related quality of life[30], therefore, more attention should be paid to the prevention and treatment of periodontal disease.

Previous studies have shown proper brushing, and appropriate use of dental care products[31, 32]. Brushing is the basic method to maintain oral health, as it can remove supragingival plaque on the facial and lingual/palatal surfaces, our survey shows that 92.9% of patients could brush their teeth twice a day or more. However, only half of the patients (53.3%) were able to rinse their mouths daily. Even fewer patients had a dental cleaning, accounting for only 26.6%. The results showed that the majority of patients had never had their teeth cleaned, probably because they had a low perception of the benefits of a dental cleaning or were not sufficiently motivated to do this, and did not receive the necessary social support[33]. A dental cleaning can effectively remove calculus, plaque, and food debris[34]. There is evidence that dental insurance plays an important role in determining dental cleanings. The availability of health insurance has a positive effect on dental cleanings. It is therefore possible to improve oral health by enrolling in a plan that offers dental insurance[35]. Studies have shown that perceived self-efficacy, perceived benefits and barriers, and fear of dentistry influence dental scaling practices[36]. The poor performance of the patient’s dental cleaning behavior indicates the need for an intervention program designed to promote dental cleaning behavior. Self-efficacy and perceived benefits of dental cleaning should be increased to reduce fears and barriers.

This study showed that 11.4% of patients had frequent gingival bleeding. Gingivitis is a common oral health problem, and untreated gingivitis can progress to periodontitis. Previous studies have shown that health-related lifestyles, such as a healthy diet, good oral health habits, and more frequent dental visits, are protective factors for the condition of the gums[37]. Gingival bleeding on brushing, as reported by self-report, is an important sentinel sign of periodontal disease, and gingival inflammation in particular. By identifying gingival bleeding on brushing, periodontal disease may be detected earlier, and better prevented and treated, thus reducing its global burden[38]. Our study shows that patients with inadequate OHL tended to rinse their mouths less and had a higher likelihood of bleeding gums. 54.3% of patients had a dental visit in the past year, much lower than the percentage of 89.5% in a Ugandan study[39].

Self-rated oral health is a comprehensive evaluation of the oral health of the participants based on their health and overall subjective feelings. This study showed that 15.2% of patients rated their oral health as poor, 69.6% rated their oral health as fair, and only 15.4% rated their oral health as good, which is much lower than a survey of Indigenous Australians[40]. As a previous survey shows, compared with Netherlands 35-44-yearolds, Chinese 35-44-year-olds rated their oral health as poor or fair more often, and Netherlands 35-44-year-olds rated their oral health as very good four times more often than Chinese[41]. Overall, the better the self-rated oral health status, the higher the oral health literacy score. Previous studies have shown that self-rated oral health was associated with oral health utilization among the Chinese population[24], patients with poorer self-rated oral health tend to have poorer oral health behaviors and oral health status[14], and overall health status also tends to be poorer, such as self-care and mobility, and oral health status is also closely related to mental health status, and patients with poorer oral health status also tend to have more severe anxiety and depression, etc.[40].

Characteristics associated with inadequate OHL were consistent with the scientific literature[22]. This study shows that economic burden is influential in oral health literacy. Patients with economic burdens tend to have inadequate OHL, which is consistent with previous studies[42].

Patients with adequate OHL were more likely to have a dental visit in the last year than patients with inadequate OHL, which has been confirmed by numerous studies[10]. OHL levels are correlated with dental service utilization, according to previous studies[20, 42]. Mialhe et al.[42] observed in Brazilian adults that the higher the OHL levels, the more frequently they had a preventive dental checkup. However, this correlation was not reflected in our study, probably due to the limited number of participants in this study, especially the small percentage of patients who came for consultation. Moreover, some of the medical checkup patients may be organized by companies or hospitals instead of coming on their own, which has been mentioned by investigators and medical institution managers. In the future, the sample size could be further expanded and the patients who underwent medical checkups or initiated consultations could be further subdivided to explore the correlation between OHL and the reasons for dental visiting.

This study shows that patients with economic pressure tend to have inadequate OHL. The results of this study support the previously established relationship between inadequate OHL and oral health behaviors including gargling and dental visiting[13, 43]. In addition, we found an association between oral health literacy and gum bleeding during tooth brushing.

Our study is a novel one in the sense that it presents OHL, oral health behavior and oral health status data in China, where studies on OHL are just beginning. In this study, a validated instrument was used to measure wider aspects of OHL, such as the ability to interpret and use oral health information and services. While earlier tools mainly assess word recognition, numeracy, and reading skills, such as Rapid Estimate of Adult Literacy in Dentistry (REALD) or Test of Functional Health Literacy in Dentistry (ToFHLiD), and because the test language is English, it is less applicable in non-English speaking countries. HeLD-14 assesses a wide range of aspects of OHL and has been validated as having good psychometric properties. This was the first research to evaluate oral health literacy in dental patients in China. The inadequate OHL in this study was associated with some oral health behaviors and oral health status. Based on the results of our study, we find that in addition to health determinants, OHL can have a significant impact on oral health outcomes, oral health behaviors, and dental services use. The significance of this fact is that OHL can be changed through health promotion strategies to positively impact oral health outcomes.

## Limitation

This study has several limitations. First, we examined oral health literacy and oral health behaviors using a cross-sectional design. As a result, we could not examine the causality relationships between the two variables. Second, self-reported questionnaires are vulnerable to recall bias. Despite such limitations, our study is one of the few in the literature to examine the association between oral health literacy, oral health behavior, and oral health status in dental patients. The results of this study may serve as an evidence base to inform future studies.

## Conclusion

The oral health literacy of dental patients is at a medium level overall. Oral health literacy among dental patients is associated with economic burden, mouthwash, gum bleeding, and dental visiting. Health care providers should consider improving oral health literacy among dental patients. OHL is an emerging research area, and critical information is still needed on its influencing factors, impact pathways, oral health outcomes, and intervention strategies. Future research can identify the influencing factors of oral health literacy and develop intervention strategies to improve oral health literacy levels and oral-related quality of life.

## Data Availability

All data produced in the present study are available upon reasonable request to the authors

## Acknowledgment

We thank the first Mobile Corps Hospital of the Chinese People’s Armed Police Force for their support and cooperation.

